# Prognosis of stage IV bone cancer based on pattern of metastasis: a retrospective survival analysis of patients from the Surveillance, Epidemiology, and End Results (SEER) database

**DOI:** 10.1101/2020.10.20.20215913

**Authors:** Talha Ayaz, Megna A. Panchbhavi, Muhammad T. Kashif, Mohammed S. Abdullah, Zahra Akhtar

## Abstract

**Background:** Prognosis in bone cancer patients with metastatic disease is believed to vary based on site/pattern of spread. In 2003, the American Joint Committee on Cancer (AJCC) incorporated this observation into the TNM Staging System by subclassifying metastatic disease into M1a or M1b. We conducted a retrospective survival analysis of patients with primary bone cancer to characterize prognosis and assess outcomes in M1a versus M1b disease.

**Methods:** The Surveillance, Epidemiology and End Results (SEER) database was searched for cases of primary bone cancer presenting with metastasis from 2010 to 2015. Cases were grouped using AJCC metastatic staging as metastasis to the lung only (M1a) or other pattern of metastasis (M1b). Overall survival and cause-specific survival were assessed using Kaplan-Meier analysis and multivariate cox regression models. Multivariate models adjusted for multiple demographic, tumor characteristic, and treatment covariates.

**Results:** Five hundred and twenty-six cases met the inclusion criteria for this study. Two hundred and forty-eight were staged as M1a, and 278 were staged as M1b. Mean follow-up time for the cohort was 18.21 months (*SD* = 16.76). Fifty percent (124 of 248) of M1a and 59.4% (165 of 278) of M1b patients had died by the end of the study. Overall (*P* = .003) and cause-specific survival (*P* = .010) times were significantly lower for M1b patients via log-rank test. Adjusted analysis showed that M1b patients had poorer overall survival (HR, 1.505; 95% CI, 1.138-1.989; *P* = .004) and cause-specific survival (HR, 1.446; 95% CI, 1.091-1.918; *P* = .010) compared to M1a patients.

**Conclusion:** Metastasis pattern is an independent predictor of survival. M1a metastatic disease tends to have a better prognosis compared to M1b. This study supports the decision of the AJCC to subclassify metastatic disease for the purposes of staging and highlights the differences in prognosis between these two patterns of disease.

## Introduction

Primary malignant bone tumors are rare malignancies, with the most common forms being osteosarcoma, chondrosarcoma or Ewing sarcoma^1^. In clinical practice, the Musculoskeletal Tumor Society’s (MSTS) approach to staging these malignancies has traditionally been preferred over the conventional American Joint Committee on Cancer’s (AJCC) TNM system. In its sixth edition manual, the AJCC revised its approach for staging bone cancers to better reflect distinct prognostic classifications.

The revised staging system included new subclassifications for metastatic disease, categorizing tumors with lung-only metastasis as M1a and those with metastasis to other distant sites as M1b^2^. The latter sites are thought to be associated with a poorer prognosis; however, this distinction and the effectiveness of the subclassification scheme are not well characterized by the current literature.

The authors of this study performed a retrospective survival analysis of patients with metastasized primary malignant bone tumors to better understand these metastatic subclassifications and characterize prognosis associated with M1a- and M1b-type metastatic disease.

## Methods

Cases of primary bone cancer with metastasis from 2010 to 2015 were identified in the Surveillance, Epidemiology and End Results (SEER) database using primary site codes C40.0-C41.9 designating ‘Bones, Joints, and Articular Cartilage.’ The specific SEER dataset used for data selection was titled: Incidence - SEER 18 Regs Custom Data (with additional treatment fields), Nov 2017 Sub (1973-2015 varying)^3^. Patients with incomplete follow-up data or unknown cause of death were not included in this study. The cohort was sub-grouped for analysis into M1a or M1b using the AJCC’s seventh edition metastasis (M) descriptor. Descriptive statistics were calculated to compare subgroup demographic, tumor, and treatment characteristics using chi-square analysis. Overall survival and cause-specific survival were assessed using log-rank test and multivariate Cox regression analysis. Cox regression models were adjusted for multiple sociodemographic, tumor characteristic, and treatment factors. Adjusted covariates included the following: age, sex (male or female), race (white, black, Asian/Pacific Islander, American Indian/Alaska native, or unknown), ethnicity (non-Hispanic or Hispanic), marital status (married, single, divorced/separated, widowed, or unknown), percent of poverty in county of residence, primary site, tumor grade (I, II, III or IV), tumor (T) site (T0, T1, T2, T3 or unknown), lymph nodes (N) affected (N0, N1 or unknown), cancer type (osteosarcoma, chondrosarcoma, Ewing tumor, or other specified and unspecified bone tumors), treatment with primary tumor surgery (yes, no, or unknown), non-primary site surgery (yes, no, or unknown), radiotherapy (yes, no, or unknown) and chemotherapy (yes, no, or unknown).

## Results

Five hundred and twenty-six cases with a mean follow-up time of 18.21 months (SD = 16.76) met the inclusion criteria for this study. Median age was 21 years (range, 2-85+ years). Two hundred and forty-eight cases were staged as M1a and 278 as M1b. Fifty percent (124 of 248) of M1a and 40.6% (165 of 278) of M1b patients were alive at end follow-up. Age (*P =* .051), sex (*P =* .419), ethnicity (*P =* .485), marital status (*P =* .242), percent of poverty in county of residence (*P =* .890), and use of chemotherapy (*P =* .259) did not significantly differ between M1a and M1b subgroups. With regard to primary site (*P* < .001), the M1b subgroup (28.8%) had a significantly higher proportion of lesions originating from the pelvis compared to the M1a subgroup (21.8%). Race (*P =* .008), T (*P* < .001), N (*P =* .015), cancer type (*P =* < .001), primary site surgery (*P* < .001), non-primary site surgery (*P =* .004), and radiotherapy (*P =* .005) also significantly differed between subgroups as shown (Table 1).

**Table 1:**
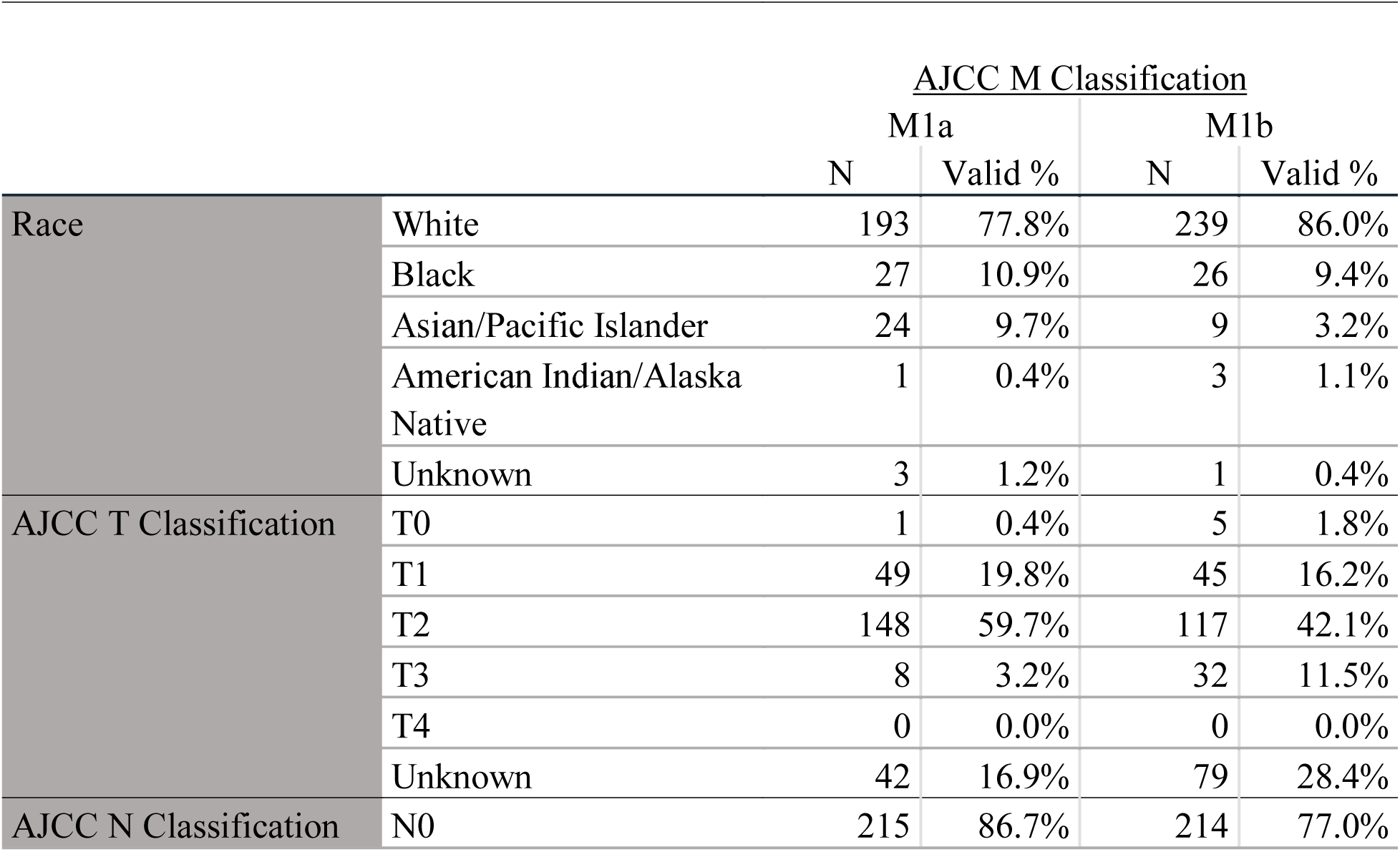
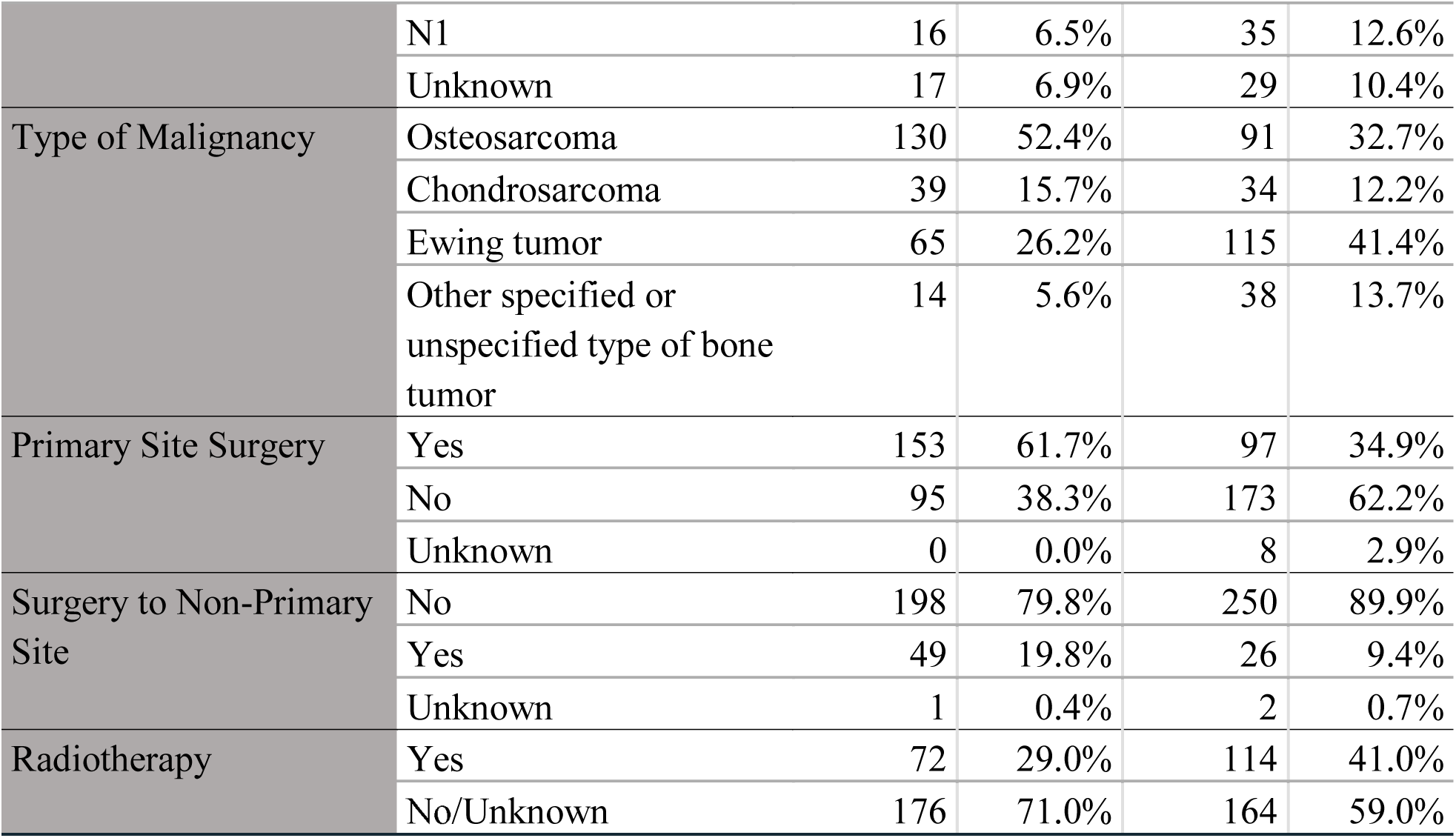
Differences in demographic, tumor and treatment characteristics of M1a and M1b Patients

Univariate log-rank analysis found that overall survival (*P =* .003) and cause-specific survival (*P =* .010) significantly differed between M1a and M1b subgroups. Mean overall survival time (*P =* .003) was 34.85 months (SE 2.044, 95% CI 30.844-38.856) for M1a patients and 27.18 months (SE 1.783, 95% CI 23.685-30.674) for M1b patients. The mean cause-specific survival time (*P =* .003) was 35.087 months (SE 2.054, 95% CI 31.061-39.112) for M1a patients and 28.181 months (SE 1.829, 95% CI 24.595-31.767) for M1b patients.

Multivariate Cox regression analysis found that M1b patients had a significantly higher all-cause (HR, 1.505; 95% CI, 1.138-1.989; *P =* .004) and cause-specific (HR, 1.446; 95% CI, 1.091-1.918; *P =* .010) mortality risk compared to M1a patients. Cumulative overall (Figure 1) and cause-specific (Figure 2) survival function curves are shown. Cox regression models were adjusted for the following covariates: age, race, sex, ethnicity, marital status, percent poverty in county of residence, grade, primary site, AJCC T classification, AJCC N classification, type of malignancy, treatment with primary site surgery, surgery to non-primary site, radiotherapy, and chemotherapy.

**Figure 1:**
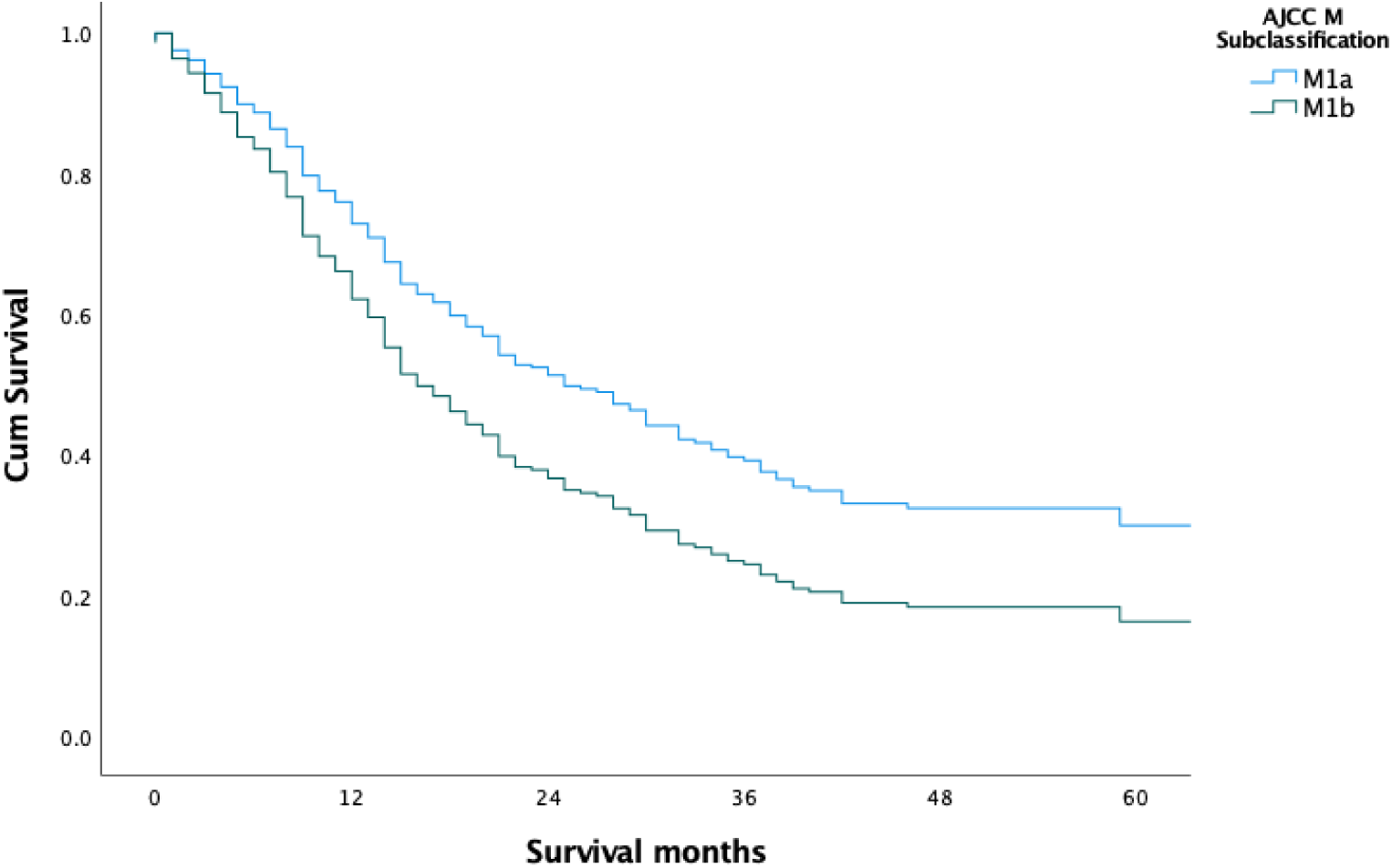
5-Year Overall Cumulative Survival Curve Based on AJCC Metastatic (M) Subclassification.

**Figure 2:**
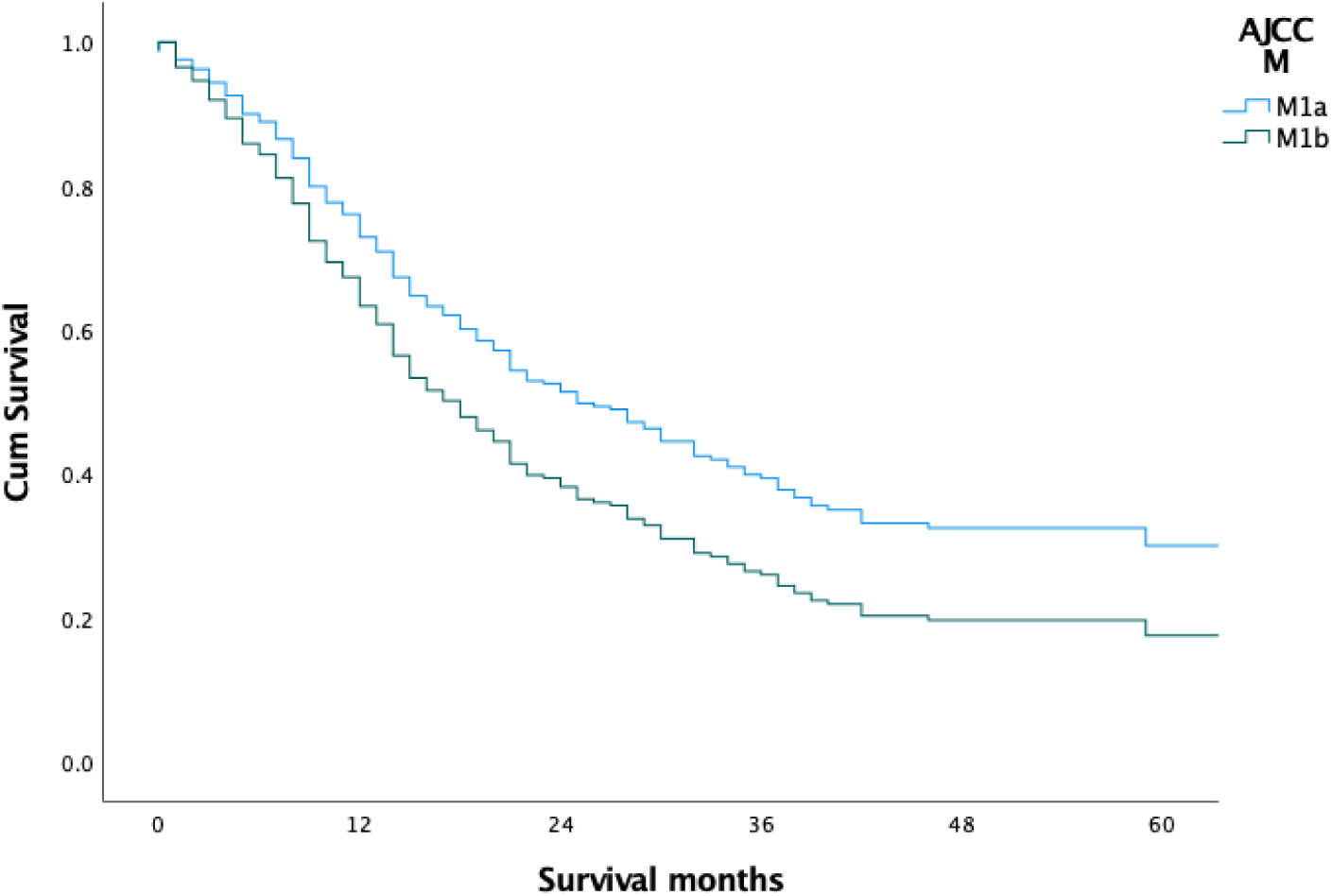
5-Year Cause-Specific Cumulative Survival Curve Based on AJCC Metastatic (M) Subclassification.

## Discussion

Using the SEER database, we retrospectively analyzed survival in patients with metastasized primary malignancies of the bone based on AJCC subclassifications of the M1 tumor descriptor used for staging. This study contextualizes the prognostic implications and comparative outcomes of M1a and M1b metastatic disease from primary bone cancer and provides insightful evidence that supports the AJCC’s decision to subclassify the M descriptor for these malignancies. These are a rarer group of malignancies; consequently, the corresponding scientific literature lacks substantive clinical descriptions regarding specific sarcomatous lesions and the nuanced expectations in clinical course based on extent/pattern of disease. Upon covariate adjustment for sociodemographic, tumor characteristic, and treatment factors, M1b patients had significantly higher overall and cause-specific mortality rates compared to M1a patients.

Although the difference in prognosis for M1a- and M1b-type malignancies of the bone have not been specifically investigated, some studies have shown that prognosis may be better for those with lung metastasis, as these lesions can more often be resected through surgical metastasectomy^4,5,6^. Salah et al. evaluated the impact of pulmonary metastasectomy on survival of osteosarcoma patients with resectable lung metastases^4^. In the group of patients with osteosarcoma, lower mortality was reported among patients treated with metastasectomy compared to those managed with systemic therapy or those with unresectable metastases. The study ultimately found that pulmonary metastasectomy was associated with an improved survival of osteosarcoma patients with resectable lung metastases^4^. Kim et al. also found an association with pulmonary metastasectomy and prolonged survival, and Bacci et al. demonstrated that osteosarcoma patients with extrapulmonary metastases had a higher survival rate if they were treated with a combination of neoadjuvant chemotherapy, simultaneous resection of primary (and, when feasible, secondary) lesions, followed by chemotherapy^5, 6^. Since M1a osteosarcoma can be treated with pulmonary metastasectomy, this treatment may have improved the prognosis of M1a patients compared to M1b patients.

Smaller studies in the past have similarly assessed survival in sarcoma patients with metastatic disease in the lungs compared to patients with metastatic disease elsewhere^7,8,9^. Meyers et al. noted that survival for patients with osteogenic sarcoma correlated with the location of metastatic disease^7^. In a study reviewing patients with osteosarcoma, Ferguson et al. noted that only 1 of 17 patients with unresectable disease or distant bone metastases survived, in contrast to 6 of 17 patients with the lung as their only metastatic site and 2 of 3 patients with resected regional bone metastases^8^. Mialou et al. also noted that none of the osteosarcoma patients in their study presenting with only extrapulmonary metastases without any pulmonary metastases survived^9^. A handful of other studies investigating general prognostic factors in sarcoma also endorsed this pattern in their findings^10,11,12^. These aforementioned reports all found that the survival rate for patients with extrapulmonary metastases was lower than the survival rate for patients with intrapulmonary metastases, and these findings are corroborated by the current study. Only one prior investigation showed no significant difference in survival; however, this analysis addressed differences based on overall disease stage rather than analyzing metastatic disease subclassifications specifically^2^.

To the author’s knowledge, this study is the first one to compare survival outcomes for M1a and M1b osteosarcoma patients. Although several studies have reviewed osteosarcoma patients with pulmonary metastases, only a few have reviewed osteosarcoma patients with extrapulmonary metastases. This study focused specifically on M1a and M1b osteosarcoma patients, and the difference in prognosis between the two. The difference in prognosis found in this study supports the AJCC classifying system, which is based on prognosis. With growing pressure to adopt the AJCC staging system, knowing the significance of this system on the patient’s prognosis is valuable. The additional prognostic information for either M1a or M1b osteosarcoma can also help guide clinical decisions, as the survival rate may affect the patient’s goals.

This investigation encountered a number of limitations. Given that this was a retrospective analysis of patients reported within a national database, selection bias may be considered a potential limiting factor. Additionally, the use of a database naturally limited the authors to the variables and outputs made available therein. Relevant characterizing information, therefore, such as type of malignancy or method of treatment, was not available for some patients or was reported in insufficient detail. For example, some malignancies were only identifiable as ‘other specified or unspecified bone tumors’ rather than a specific histologic subtype of malignancy and employed treatment modalities were reported in a ‘yes’ or ‘no/unknown’ format. To counter this limitation, the authors extracted data for several covariates, allowing for more complete patient descriptions and subsequent control for confounders. To add to this, only cases with complete follow-up and survival data were included. Patient demographic descriptors were also limited to those made available within the database and did not include individual socioeconomic data, hence the use of a county-level descriptor to account generally for potential socioeconomic differences. Lastly, the authors could not account for miscoded data within the database resulting from human error.

## Conclusion

To our knowledge, this retrospective analysis of bone cancer patients is the first to describe and comparatively assess survival for distinct subclasses of metastatic disease defined by the AJCC staging system. This study showed that M1a metastatic disease (intrapulmonary metastasis only) tends to have a better prognosis compared to M1b metastatic disease (extrapulmonary metastasis present). This corroborates the decision to differentiate the two patterns for the purposes of characterizing disease severity and expected clinical course. Further research is necessary to help identify and subsequently address obstacles to effective treatment for bone cancer patients with distinct manifestations of metastatic disease.

## Data Availability

The data that support the findings of this study are openly available within the Surveillance Epidemiology and End Results (SEER) national cancer database. The Registry of Research Data Repositories persistent identifier for this resource is RRID: nif-0000-21366. The data can be extracted through a case listing session from the dataset titled: Incidence - SEER 18 Regs Custom Data (with additional treatment fields), Nov 2017 Sub (1973-2015 varying).

http://www.seer.cancer.gov/

## Declaration of Competing Interests

The authors have declared no competing interest.

## Financial Disclosure

This study did not receive any funding or financial support.

## Ethical Statement

This study did not qualify as human subject research, as defined by the institutional IRB regulations at 45 CFR46.012 (d)(f) and at 21 CFR 56, as it was conducted using de-identified data from a publicly accessible database, and therefore did not require IRB approval or oversight. Acknowledgements:

We would like to thank the National Cancer Institute for its establishment of the SEER program as means to promote cancer research - Surveillance Research Program, National Cancer Institute SEER*Stat software (seer.cancer.gov/seerstat) version 8.3.5.

## Notes

### Author Declarations

This study did not qualify as human subject research, as defined by the institutional IRB regulations at 45 CFR46.012 (d)(f) and at 21 CFR 56, as it was conducted using de-identified data from a publicly accessible database, and therefore did not require IRB approval or oversight.

## References

1. von Eisenhart-Rothe R, Toepfer A, Salzmann M, Schauwecker J, Gollwitzer H, Rechl H: Primär maligne Knochentumoren. Der Orthopäde. 2011, 40:1121-42. 10.1007/s00132-011-1866-7

2. Heck RK, Jr., Stacy GS, Flaherty MJ, Montag AG, Peabody TD, Simon MA: A comparison study of staging systems for bone sarcomas. Clin Orthop Relat Res. 2003:64-71. 10.1097/01.blo.0000093898.12372.6c

3. Surveillance, Epidemiology, and End Results (SEER) Program (2018). Accessed: October 1, 2020: www.seer.cancer.gov.

4. Salah S, Ahmad R, Sultan I, Yaser S, Shehadeh A: Osteosarcoma with metastasis at initial diagnosis: Current outcomes and prognostic factors in the context of a comprehensive cancer center. Mol Clin Oncol. 2014, 2:811-16. 10.3892/mco.2014.325

5. Kim S, Ott HC, Wright CD, et al.: Pulmonary resection of metastatic sarcoma: prognostic factors associated with improved outcomes. Ann Thorac Surg. 2011, 92:1780-6; discussion 86-7. 10.1016/j.athoracsur.2011.05.081

6. Bacci G, Rocca M, Salone M, et al.: High grade osteosarcoma of the extremities with lung metastases at presentation: treatment with neoadjuvant chemotherapy and simultaneous resection of primary and metastatic lesions. J Surg Oncol. 2008, 98:415-20. 10.1002/jso.21140

7. Meyers PA, Heller G, Healey JH, et al.: Osteogenic sarcoma with clinically detectable metastasis at initial presentation. J Clin Oncol. 1993, 11:449-53. 10.1200/jco.1993.11.3.449

8. Ferguson WS, Harris MB, Goorin AM, et al.: Presurgical window of carboplatin and surgery and multidrug chemotherapy for the treatment of newly diagnosed metastatic or unresectable osteosarcoma: Pediatric Oncology Group Trial. J Pediatr Hematol Oncol. 2001, 23:340-8. 10.1097/00043426-200108000-00004

9. Mialou V, Philip T, Kalifa C, et al.: Metastatic osteosarcoma at diagnosis: prognostic factors and long-term outcome--the French pediatric experience. Cancer. 2005, 104:1100-9. 10.1002/cncr.21263

10. Cotterill SJ, Ahrens S, Paulussen M, et al.: Prognostic factors in Ewing’s tumor of bone: analysis of 975 patients from the European Intergroup Cooperative Ewing’s Sarcoma Study Group. J Clin Oncol. 2000, 18:3108-14. 10.1200/jco.2000.18.17.3108

11. Wuisman P, Enneking WF: [Staging of osteosarcoma with skip metastases]. Z Orthop Ihre Grenzgeb. 1990, 128:457-62. 10.1055/s-2008-1039596

12. Bielack SS, Kempf-Bielack B, Delling G, et al.: Prognostic factors in high-grade osteosarcoma of the extremities or trunk: an analysis of 1,702 patients treated on neoadjuvant cooperative osteosarcoma study group protocols. J Clin Oncol. 2002, 20:776-90. 10.1200/jco.2002.20.3.776

